# Evaluation of a novel direct capture method for virus concentration n wastewater from COVID-19 infectious ward and correlation analysis with the number of inpatients

**DOI:** 10.1101/2021.11.17.21266445

**Authors:** Manami Inaba, Ryohei Nakao, Fumiko Imamura, Yutaka Nakashima, Seiji Miyazono, Yoshihisa Akamatsu

**Affiliations:** Graduate School of Science and Technology for Innovation, Yamaguchi University, 2-16-1, Tokiwadai, Ube, Yamaguchi, 755-8611, Japan; NIPPON KOEI CO., Research & Development Center Center for Advanced Research, 2304 Inarihara,Tsukuba, Ibaraki, 300-1259, Japan; Yamaguchi Prefectural Grand Medical Center, 77, Osaki, Hofu, Yamaguchi, 747-8511, Japan

**Keywords:** Virus concentration, Direct capture, PEG precipitation, Wastewater-based epidemiology, Wastewater, SARS-CoV-2

## Abstract

The global outbreak of the SARS-CoV-2 pandemic has increased the focus of Wastewater-based epidemiology (WBE) studies as a tool for understanding the epidemic and risk management. A highly sensitive and rapid method for the virus concentration from wastewater is needed to obtain the accurate information for early detection of SARS-CoV-2 outbreak and epidemic. In this study, we evaluated the efficiency of the direct capture method provided from Promega, based on column adsorption using the wastewater from actual infectious diseases ward. The efficiency of the nucleic acid extraction-purification process was also evaluated by Maxwell® RSC instrument (fully automated extraction) and QIAamp Viral RNA mini kit (manual extraction). The obtained SARS-CoV-2 data from wastewater were analyzed with the number of inpatients which is the consideration of the severity and the days of onset. The combination of direct capture and Maxwell’s method (DC-MW) was suggested to be a highly sensitive and simple method with better concentration efficiency and quantification than other methods. Moreover, the inpatient conditions (severity and days of after onset) should be considered to accurately understand the actual status of the correlation between the number of inpatients and SARS-CoV-2 concentration in wastewater. The highly sensitive method of DC-MW was suggested to assess more actual situation of SARS-CoV-2 shedding into the wastewater.

## INTRODUCTION

Wastewater-based epidemiology (WBE) is an effective way to detect and quantify specific substances (e.g., pathogenic microorganisms, drugs, pesticides, etc.) in wastewater and to collect information on behavior (e.g., prevalence, consumption, etc.) in the catchment area of the target substance (Victoria et al., 2016, Salvatore et al., 2015. Prevost et al., 2015). Previous WBE studies have focused the early detection of epidemics in enteric viruses such as norovirus and the WHO poliovirus eradication program (WHO 2015, Yoshida et al., 2000, Kazama et al., 2016). The WBE is also expected to be utilized in the novel coronavirus disease (COVID-19) caused by severe acute respiratory syndrome coronavirus 2 (SARS-CoV-2) infections, which are still pandemic. SARS-CoV-2 has been identified to be shed in the feces as well as in the sputum and swabs of patients respiratory (Wolfel et al., 2020). The detection of SARS-CoV-2 in the wastewater has also been reported in many countries (Haramoto et al., 2020, Wu et al., 2020a, Randazzo et al., 2020, Bar- Or et al., 2020). A disturbing characteristic of the coronaviruses is the presence of many asymptomatic patients, who shed coronaviruses as well as symptomatic patients (Avanzato et al., 2020, Polo et al., 2020). Furthermore, it has been estimated that the period of coronavirus shedding from infected individuals begins several days (3-7 days) before the onset of the infection, peaks around the day of onset, and lasts until several days (3-7 days) later (He et al., 2020). These indicate that SARS-CoV-2 shed from patient can exist in the environment even before the actual patients are confirmed. SARS-CoV-2 had been detected in wastewater samples around the world before the virus was detected in the population of the study area (Ahmed et al., 2020, La Rosa et al., 2020). Thus, WBE is expected to be an effective tool for the early detection of SARS-CoV-2 and the warning of the epidemic, spread, and infection risks.

The WBE process for virus can be roughly divided into four processes (wastewater sample collection, virus concentration, extraction and purification of the viral genome, and genome quantification). The virus concentration is particularly essential and important process to increase the sensitivity of virus detection because viruses in public wastewater are often in low concentrations. Various methods such as polyethylene glycol precipitation (PEG), ultracentrifugation, ultrafiltration (UF), and negatively charged membrane method have been applied to concentrate the wastewater effluent (Kazama et al., 2016, Huang et al., 2000, Katayama et al., 2002, Torii et al., 2020). Among these, the PEG is broadly used in WBE for SARS-CoV-2 because it is representative and inexpensive (Hata et al., 2020, Farkas et al., 2021, Kocamemi et al, 2020., Graham et al., 2021, D’Aoust et al., 2021). Although the efficiency of concentration by PEG is not high and stable, it has been known that the efficiency varies with the sample condition and the operator skill (Salvo et al. 2021, Ahamed et al., 2020, Hata et al., 2021, Torii et al., 2022). In addition, the addition PEG and NaCl amounts, incubation time, and centrifugation conditions vary with study, and no standardized recommended conditions have been determined (Wu et al., 2020b, Lewis and Metcalf 1988, Kocamemi et al. 2020, Barril et al., 2021). Thus, it is necessary to establish a highly efficient and simple method for quantifying the virus concentration from wastewater.

The WBE has increased attention due to the COVID-19 epidemic, and various products for virus concentration in wastewater have been provided. The Viral RNA/DNA Concentration and Extraction Kits for Wastewater (Direct capture kit, DC) released by Promega is a column-based aspiration method for the direct capture and concentration of total nucleic acids (TNA) in wastewater. Promega reported that the concentration of viral nucleic acids is faster than that of the PEG and the stable recovery rate is achieved (Mondal et al., 2021). In addition, it has the advantage of reducing the burden on the operator and ensuring safety, because it takes only about two hours of work, and the initial process includes inactivation process by isopropanol and protease. The DC method is expected to be a highly efficient, simple, and stable method for virus concentration in wastewater.

The specific objective of this study is to evaluate the differences in concentration efficiency between the conventional PEG and the DC method by detecting and quantifying the SARA-CoV-2 in actual wastewater.

## MATERIALS AND METHODS

### Sample collection

Wastewater samples were collected from four sites. One of the four sites was an independent septic tank located at the infectious disease ward in the general hospital in the Yamaguchi Prefecture. A total of seven samples were collected at this medical wastewater (MedWW) site from May 1 to June 17 in 2021. The other three sites were in the public wastewater treatment plants (WWTPs: -A, -B and -C), and one sample per site was collected on June 4 in 2021. The water quality items (water temperature, pH, electrical conductivity (EC), total dissolved solids (TDS), and salinity) measured in the each collected water sample are summarized in Table 1. All samples collected into sterile bottles were transported on ice to the laboratory and stored at -80°C until the experimental treatment.

**Table 1.** Water quality data of the collected wastewater samples.

| Site | Date | Temperature<br>(°C) | pH | EC<br>(μS) | TDS<br>(ppm) | Salinity<br>(ppt) |
| --- | --- | --- | --- | --- | --- | --- |
| MedWW | May 1 | 18.8 | 7.42 | 371 | 264 | 0.18 |
|  | May 7 | 19.6 | 6.16 | 760 | 522 | 0.37 |
|  | May 13 | 20.9 | 6.23 | 788 | 560 | 0.39 |
|  | May 20 | 23.1 | 9.44 | 222 | 158 | 0.11 |
|  | May 27 | 19.6 | 7.04 | 650 | 461 | 0.33 |
|  | June 3 | 22.3 | 7.09 | 805 | 568 | 0.40 |
|  | June17 | 25.8 | 7.25 | 380 | 261 | 0.19 |
| WWTP-A | June 4 | 21.5 | 7.33 | 359 | 253 | 0.18 |
| WWTP-B | June 4 | 22.7 | 7.12 | 301 | 215 | 0.15 |
| WWTP-C | June 4 | 21.9 | 7.25 | 516 | 369 | 0.26 |

### Concentration methods of Virus and Total nucleic acid from wastewater samples

The 40 mL subsample of each collected wastewater was conducted for concentrating the virus and TNA by PEG or DC method, respectively (Fig.1). Each experimental condition was carried out in triplicate. Bacteriophage φ6 (NBRC 105899, NITE) which replicates in *Pseudomonas syringae* (NBRC14084, NITE) as a host was added to the 40 mL subsample to use it as whole process control (WPC) (Haramoto et al. 2018) of monitoring for the efficacy of virus concentration - quantitative reverse-trancecription PCR (qRT-PCR) processes. Wastewater subsamples which spiked φ6 were incubated at 4°C for 1 to 2 hr to reach the liquid– solid partitioning at equilibrium, followed by the virus concentration (Torii et al., 2021).

**Fig.1.**
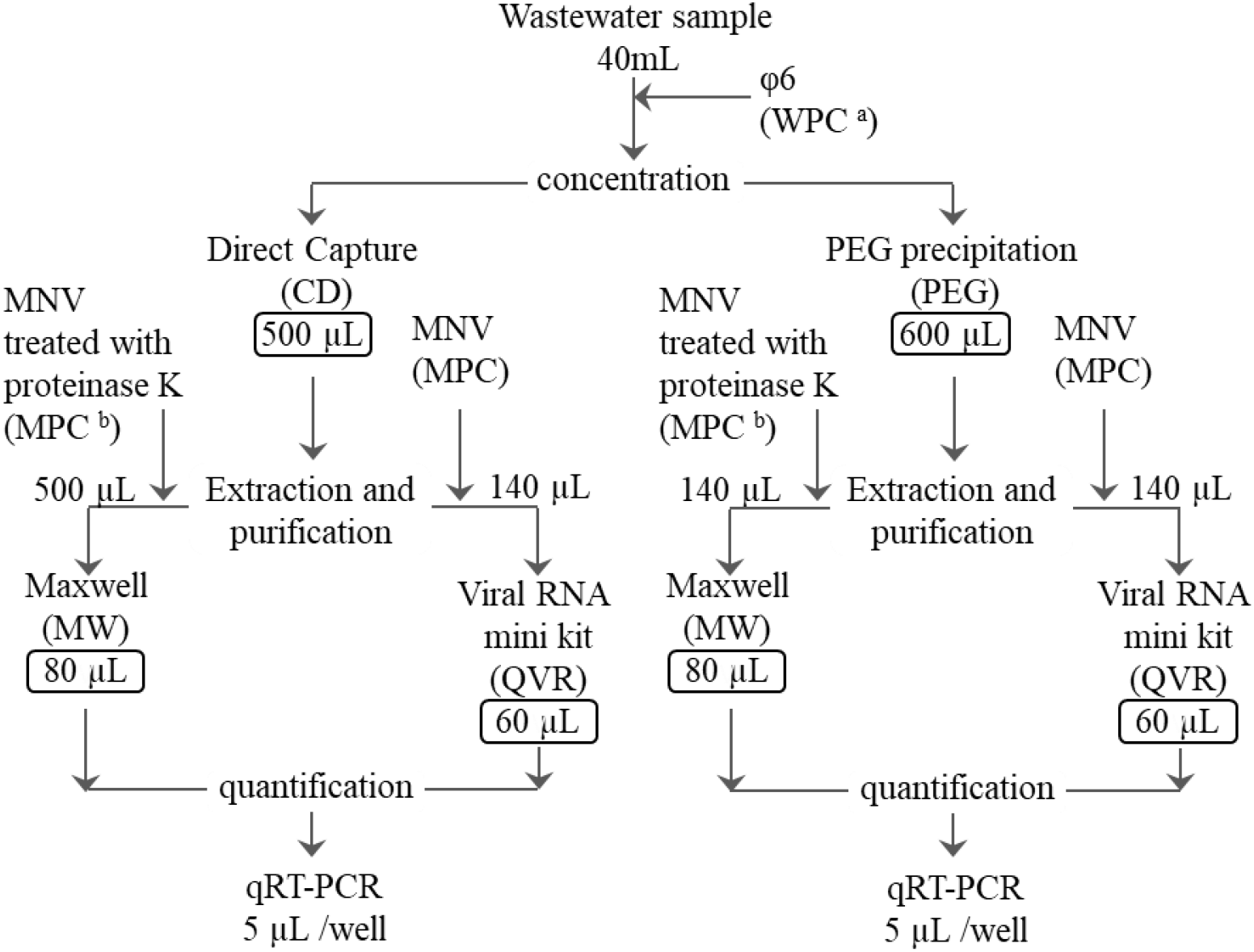
Workflow of wastewater concentration and extraction-purification processes in each comparison experiment method. Values in the squares indicate the recovered products volume. Values without a square indicate amount of specimen. ^a^ WPC: Whole process control. ^b^ MPC: Molecular process control.

Polyethylene glycol precipitation method for virus concentration was referred to Hata et al. (2021) with some modifications. Briefly, before the subsample was centrifuged at 3000 g for 5min to remove the large substance, the recovered supernatant was mixed with 4.0 g of PEG8000 and 2.35 g of NaCl at a final concentration of 10 % (w/v) and 1.0 M, respectively. The mixture was incubated on the shaker at 4 °C overnight. Then, the mixture was centrifuged at 5350 g for 70 min. The pellet was resuspended with 600 µL phosphate buffer. These virus concentrated solutions were stored at -80 °C until the molecular process.

The direct capture method to concentrate the TNA was performed in accordance with the manual provided by Promega. The subsample which reached the liquid– solid partitioning at equilibrium was treated with 500 µL of supplied protease solution under at RT for 30 min. The treated subsample was centrifuged at 3000 g for 10 min to remove the large substances. The supernatant which divided to about 20 mL into two tubes added 6 mL of binding buffer 1 and 500 µL of binding buffer 2. Additionally, 24 ml of isopropanol was added, and then mixed well by inversion. The sample mixture was poured into the PureYield™ Midi Binding Column, and applied the vacuum to capture the TNA in sample on the column. As pretreatment prior to elution, 5 mL inhibitor removal wash solution pass through the column, following to 20 mL of RNA wash solution. The column was placed in the Eluator™ device with set 1.5 mL tube and conducted to the elution process. TNA including virus genome was eluted from the column with 250 µL of pre-heated (60 °C) nuclease-free water, which was repeated twice. The TNA except for a few samples stored at -80°C until next molecular process was immediately subjected to RNA extraction-purification.

### Viral RNA extraction and purification

Samples concentrated by PEG and DC method were subjected to RNA extraction-purification using a QIAamp Viral RNA mini kit (QVR, Qiagen) and Maxwell® RSC instrument (MW, Promega), respectively. Before initiating the molecular process, Murine norovirus (MNV) was spiked to the sample as a molecular process control (MPC) (Rachmadi et al. 2021, Torii et al. 2021) for the RNA extraction-purification and qPT-PCR processes. The concentration of spiked MNV was 1.2 × 10^5^ – 3.0 × 10^6^ copies (10 µL MNV solution), which was spiked for concentrated products obtained from each method. In this regard, the MNV spiked for the concentrated sample subjecting to RNA purification using MW was treated by 0.1x volume of protease K (20mg/mL, Qiagen) at 56 °C for 1 hr, prior to spiked. 140 µL of the concentrated product obtained from each method was subjected to RNA extraction-purification using a QVR. 500 µL and 140 µL of the TNA concentrates produced by DC method and PEG, respectively, were purified by MW. The process in accordance with the manufacture protocol yielded 60 µL and 80 µL of purified RNA extraction product for QVR and MW, respectively. All RNA products were immediately applied to qRT-PCR templates for the quantification of SARS-CoV-2, φ6 and MNV, except for a few samples. TNA extracted by DC method which collected at MedWW and WWTP-A from May 7 to June 17 were once stored at - 80°C until purification process by QVR.

### qRT-PCR

The concentration of RNA from SARS-CoV-2, φ6 and MNV were determined with qRT-PCR using a StepOnePlus™ system (Thermo Fisher Scientific) according to previous reports (CDC 2020, Haramoto et al. 2018, Kiajima et al. 2008). Briefly, the reaction mixture for the quantification of target viruses was prepared in a final volume of 20 µL, containing a 5 µL of TaqMan™ Fast Virus 1-Step Master Mix (Thermo Fisher Scientific), forward and reverse primer set and probe in a final concentration of 0.9 µM each and 0.125 µM, respectively, nuclease-free water and 5 µL of extracted RNA. The primer pairs and probes used previously have been described in the centers for Disease Control and Prevention (CDC (2020)), Kitajima et al. (2008) and Gendron et al. (2010). The amplification profile was conducted with one-step of RT reaction and inactivation at 50 °C for 5 min and 95 °C for 20 sec, following with 50 cycles at 95 °C for 15 s and specific annealing/extension temperature for 1 min. Each of temperature for annealing/extension was 55 °C for SARS, 60 °C for φ6, and 56 °C for MNV. All reactions were performed in triplicate.

Plasmid or oligo DNA of target sequence for standard curve was prepared at 10^1^ to 10^5^ copies/well. The standard control was reacted on same plate together with the samples to obtain the standard curve. The each qRT-PCR assay showed the efficiencies of 99.9-107.3 % for SARS-CoV-2, 106.1-119.3 % for φ6, and 100.3-111.2 % for MNV by the standard curve. The correlation coefficients (r^2^) of the standard curves ranged from 0.992 to 0.998 in all target viruses.

## RESULTS AND DISCUSSION

### Efficacy of virus detection process

The recovery efficiency of φ6 as a WPC is shown in Table 2. The φ6 was added to all 10 sewage samples as an evaluation tool for the whole process from concentration to quantification by qRT-PCR to compare the efficiency of the four methods. The recovery rates of φ6 in each method, i.e., the recovery rates of virus in WPC, were DC-MW (101.4%), PEG-MW (82.7%), PEG-QVR (50.3%), and DC-QVR (21.5%), in descending order in terms of the geometric mean (GM). In DC-MW, PEG-MW, and PEG-QVR methods, the recovery rates of samples except for the MedWW sample on May 27 processed by PEG-QVR were more than 10%. Seven samples in DC-QVR method were preserved under -80°C after the concentration-recovery of RNA with DC. The recovery rates of these seven samples were 5.07∼23.4% (GM =11.9%). In addition, the recovery rates of the two samples (May 13 and 20, MedWW) were less than 10%. On the other hand, the GM of the recovery rate of φ6 on the other three samples was 89.3%. Under the DC-QVR method, comparing the group which was once subjected to the freeze and thaw process (seven samples collected at MedWW and WWTP-A from May 7 to June 17) and the other group which w/o freeze and thaw process, the recovery rate between the groups were significantly different (t-test, p < 0.05). It suggests that the freeze and thaw process could damage the recovered nucleic acid.

**Table 2.**
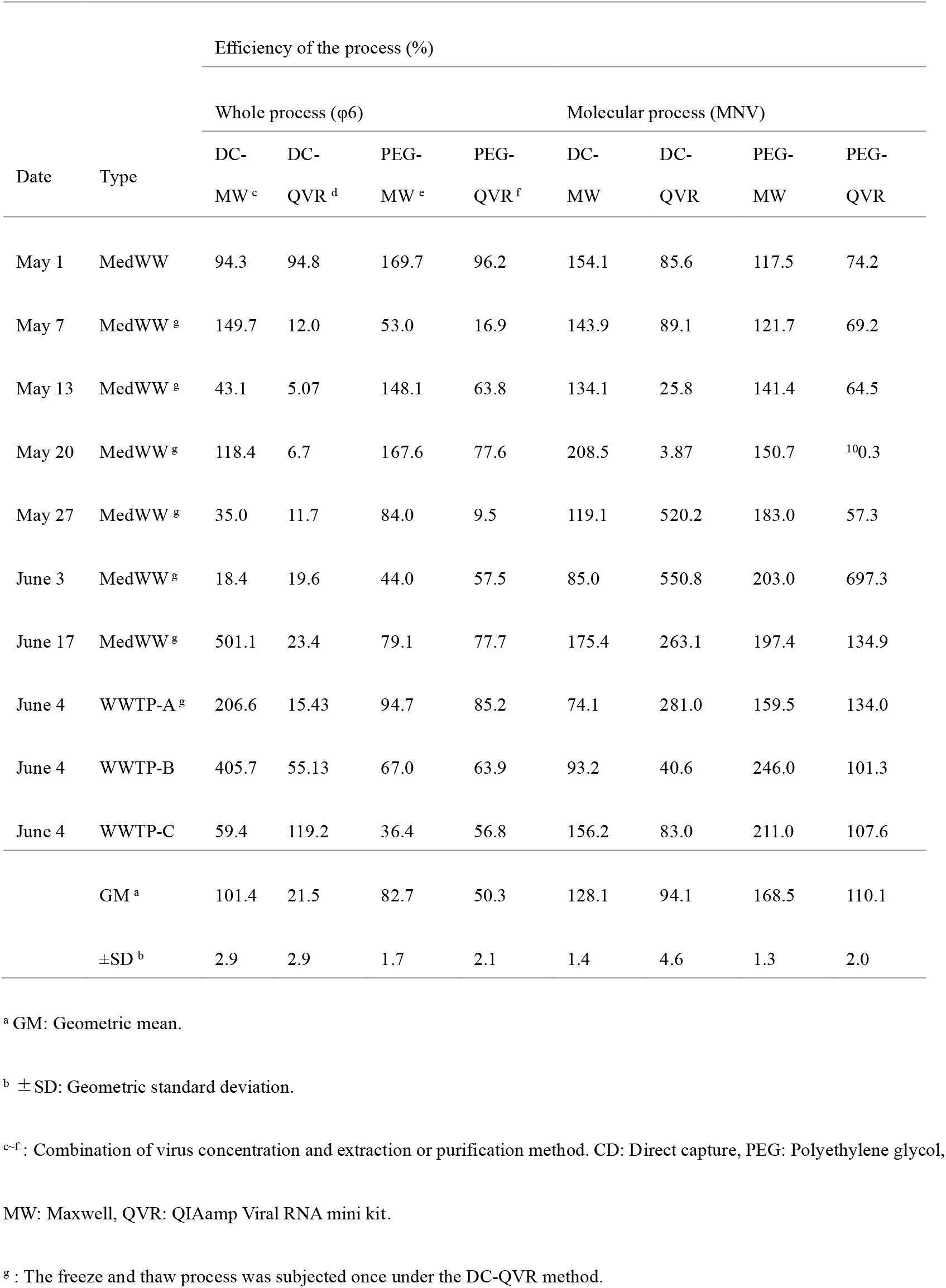
Efficiency of each process in four methods based on WPC and MPC quantitative analyses.

The efficiencies of DC and PEG in the RNA extraction-purification and qPT-PCR processes measured by spiked MNV for MPC with MW were 128.1% and 168.5% in terms of geometric mean, and their geometric standard deviation were 1.4 and 1.3, respectively. The efficiencies of DC and PEG for MPC with QVR were 94.1% and 110.1% in terms of geometric mean, and their geometric standard deviation were 4.6 and 2.0, respectively. Except for one sample (MedWW May 20, DC-QVR), the efficiency of the molecular process exceeded 10%. In the report of virus quantification from water environments, if the recovery rate of MPC is less than 10%, the quantification is evaluated to be inhibited by the present of foreign substances in the sample (Hata et al., 2021, Hata et al., 2017). Since the MPC of the same sample treated by the other three methods was above 10%, it is not considered to be affected by inhibition. On the sample of which efficiency was less than 10% processed with DC-QVR (May 20, MedWW), WPC was also low, suggesting that the actual processes of concentration and recovery were not successfully operated.

The efficiency of virus concentration has been evaluated by adding φ6 as WPC. The PEG concentration efficiencies in this study were 82.7% and 50.3% with geometric standard deviations 1.7 and 2.1 for PEG-DC and PEG-QVR, respectively, showing relatively stable and high performances. Previous studies have shown that the virus concentration rates from wastewater treated with PEG were as low as less than 10% in some cases (Salvo et al. 2021, Philo et al., 2021, Barril et al. 2021). In addition, there is no unified or standardized method for the concentration of the additive PEG, pretreatment, and centrifugation time and g (Flood et al., 2021, Sharma et al., 2021, Barril et al., 2021, Torii et al., 2022). It had been also suggested in previous reports that there are differences in the recovery efficiency depending on the target virus (Haramoto et al., 2018, Torii et al., 2021, Pérez-Cataluña et al., 2021). The concentration efficiency was thought to vary significantly with the reported cases and workers. Therefore, although virus concentration by the PEG was a relatively low-cost method and has been applied in many studies (Salvo et al., 2021, LaTurner et al., 2021), in practice, it was necessary to select the optimized conditions depending on the target virus and the condition of the wastewater sample.

Furthermore, the virus concentrates recovered by PEG method remain infectious, although this is one of advantages on this method. The virus concentration by the PEG method possessed an infection risk to the workers who handle it, and there were restrictions on the facilities. In contrast, the process of virus concentration from the wastewater by DC was conducted for the degradation at the initial step by protease, and could minimize the risk of infection to the workers. The process following the enzymatic treatment was simple (i.e., involving centrifugation, mixing of reagent buffers, and passing through a column) and were not likely to cause efficiency biases by the operators. The operation time of DC method according to the recommended protocol was about 1 hr for 40 mL of wastewater (see. https://www.promega.com/products/nucleic-acid-extraction/viral-rna-extraction-viral-dna-extraction/wastewater-viral-rna-dnaextraction/?catNum=A2991#protocols). In the DC method, the adsorbed TNA on column had been washed with the attached buffer before the elution and recovery process. This process was thought to be effective in the molecular process because it removes substances that inhibit the subsequent process, such as proteins. In our study, nucleic acid recovery and purification efficiencies were high for both MW and QVR; however, MW was found to be more efficient and stable. The sample volume that could be tested for MW was 500 µL, while that for QRV was 140 µL. The extracted nucleic acids recovered by each method were 80 µL and 60 µL, and the concentration ratio was 6.3-fold and 2.3-fold, respectively, with a higher ratio for MW. With regarded to the PEG-concentrated samples, in this study, 140 µL of PEG-concentrated samples were subjected to the extraction and purification processes of MW and QVR. These results showed that higher efficiency was obtained under the MW purification and recovery method (Table 2). In MW, TNA in the liquid phase was adsorbed on magnetic beads, and then the purification process such as washing was performed in another liquid phase. Therefore, more purified TNA was thought to had been obtained with less inhibitors carried in. In addition, the MW is a fully automated system, and no differences were in the skill of the operators. The conditions of stable purification by full automation and less introduction of inhibitors could have been contributed to the high efficiency and stable recovery and purification rate.

### Detection of SARS-CoV-2 from wastewater

In this study, qRT-PCR was performed using the CDC_N2 assay which region encoding the SARS-CoV-2 spike protein gene (CDC 2020). Table 3 and Fig. 2 show the results detected and quantified by each concentration and molecular method. The results in Table 3 are the quantitative data of SARS-CoV-2 per reaction well. Quantitative values were shown only for those with a concentration of higher than 1×10^1^ copies / reaction. The concentrations smaller than 10^1^ copies / reaction were marked with “+”, and those that were negative were shown as “-”. The DC-MW method could be obtained the highest number of quantitative data, 5 out of 10 samples. The number of samples quantified by the other three methods were 3 out of 10. In the same sample, the concentration quantified per reaction was highest in the DC-MW method (except for MedWW May 27, DC-QVR). Furthermore, the two samples (MedWW May 20, June 17) were quantified only by DC-MW, which could not be quantified or detected by other methods. Comparing the quantitative values, it was confirmed that SARS-CoV-2 concentration of DC-MW was 1.9 to 5.2 times higher than those of the other three methods. In addition, the number of samples detected, also included below the limit of quantification (LOQ), was highest in the DC-MW method (9/10). The number of samples detected by the other three methods was 6/10 to 7/10. Quantitative values of copies per L in the wastewater are shown in Fig.2. When converted to the unit of copies per L, the quantitative value of PEG-MW tended to be higher than those of other methods. The magnification when converting the quantitative value of each method to copies/L was 400 times for DC-MW, 1080 times for DC-QVR, 1720 times for PEG-MW, and 1290 times for PEG-QVR, respectively. This magnification depended on the amount of wastewater contained in the template used per reaction. The high magnification was caused by small amount of wastewater used for qRT-PCR per reaction. The calculated amount of raw wastewater added per reaction of DC-MW was 2.5 mL, and it was higher than other methods (other methods were 0.58 to 0.93 mL per reaction). Thus, DC-MW is considered to have the highest quantification per reaction. Furthermore, the LOQ per L was also lowest. These indicate that the concentration-quantification by the DCMW method is most accurate and the detection sensitivity is highest among the methods in this study.

**Table 3.** Detection and quantification results of SARS-CoV-2 from wastewater by each virus concentration and extraction-purification method.

| Sample | Detection and quantification (copies/reaction)<sup>a</sup> |  |  |  |
| --- | --- | --- | --- | --- |
|  | DC-MW | DC-QVR | PEG-MW | PEG-QVR |
| MedWW May 1 | 46.2 | 23.9 | 19.8 | 18.5 |
| MedWW May 7 | 45.0 | 30.0 | 17.1 | 12.5 |
| MedWW May 13 | 125.3 | + | 24.3 | 34.9 |
| MedWW May 20 | 60.0 | + | + | + |
| MedWW May 27 | + | 10.1 | + | + |
| MedWW June 3 | + | — | + | + |
| MedWW June 17 | 10.0 | + | — | — |
| WWTP-A | — | — | — | — |
| WWTP-B | + | — | — | — |
| WWTP-C | + | — | — | + |
| Positive quantification | 5 (50.0 %) | 3 (30.0 %) | 3 (30.0 %) | 3 (30.0 %) |
| Positive detection | 9 (90.0 %) | 7 (70.0 %) | 6 (60.0%) | 7 (70.0%) |

<sup>a</sup>: Quantification data was shown only for those that had a more than $1.0 \times 10^0$ copies/reaction of the concentration. +: Positive detection. The concentration was smaller than $1.0 \times 10^0$ copies/reaction. —: Negative.

**Fig.2.**
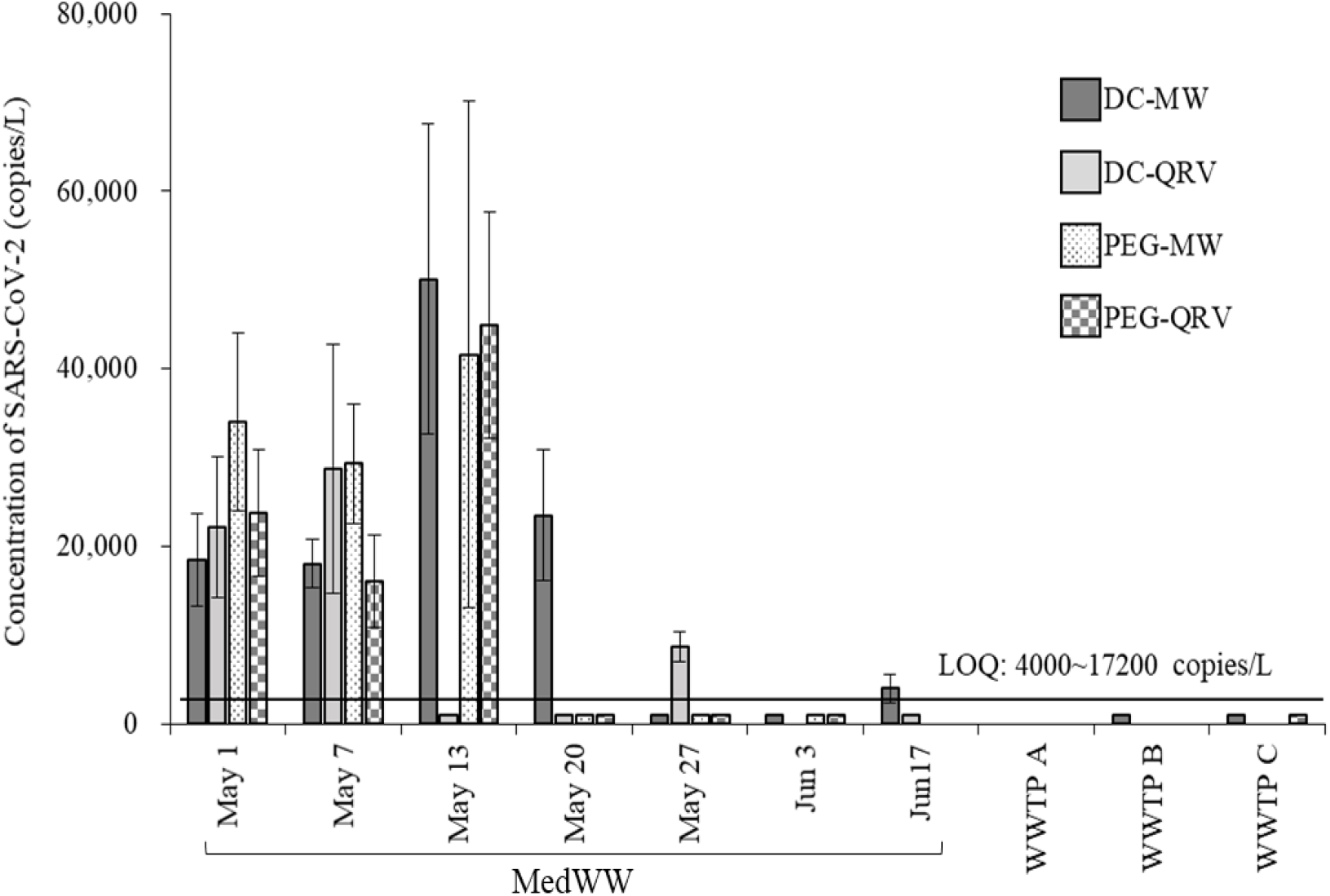
Concentrations of SARS-CoV-2 in wastewater treated by four difference types of concentration and extraction-purification methods. LOQ: Limit of Quantification. Symbols below the LOQ line indicate that virus genome were detectable, but the concentrations were smaller than 1×10^1^ copies/reaction.

### Relationship between the SARS-CoV-2 concentration in wastewater and inpatient number

The cross-correlations among the methods are shown in Table 4. The crosscorrelations among the three conditions (DC-MW, PEG-MW and PEG-QVR), which showed a positive correlation with the number of inpatients, were high. The consistency among the three conditions applied to the actual samples suggests the reliability of the obtained concentration. Fig.3 shows the correlation between the SARS-CoV-2 concentration obtained by each concentration/extraction-purification method and the number of inpatients in the infectious disease ward from which MedWW was collected. The results of Fig. 3 show that the concentrations of virus detected in the wastewater from the infectious disease ward tended to increase as the number of inpatients increased. The concentrations obtained from the three conditions other than DC-QRV showed a positive correlation with the number of inpatients. On the other hand, the concentration obtained from DC-QRV showed a negative correlation. In addition, DC-MW gave relatively higher concentration and the highest number of quantifiable samples. These suggest that DC-MW was the most reliable method for the data analysis among the methods in this study. As already discussed in “Efficacy of virus detection process” section, the results from DC-QRV imply that the extracted RNA sample were degraded by the freezing and thawing processes and the accuracy of the data is doubtful. Therefore, it was excluded in the following discussion.

**Table 4.** Cross-correlation in four difference types of the concentration and extraction-purification methods.

|  | DC-MW | DC-QVR | PEG-MW | PEG-QVR |
| --- | --- | --- | --- | --- |
| DC-MW | 1 | 0.24 | 0.76 | 0.87 |
| DC-QVR |  | 1 | 0.70 | 0.43 |
| PEG-MW |  |  | 1 | 0.43 |
| PEG-QVR |  |  |  | 1 |

**Fig.3.**
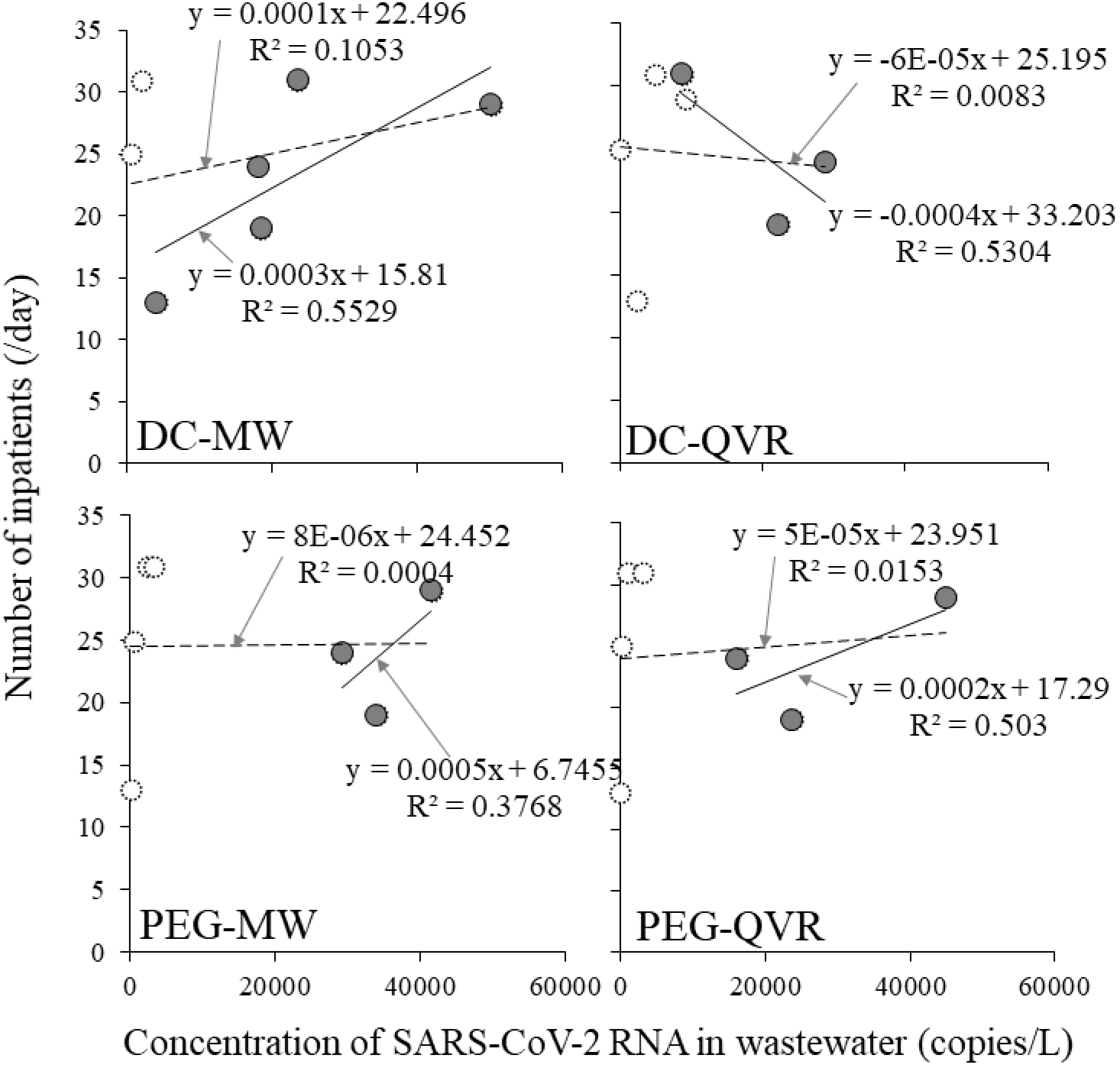
Relationship between the concentration of SARS-CoV-2 in wastewater from infectious diseases ward and the inpatient number. Gray circles indicate the relationship between the total number of inpatients and the SARS-CoV-2 concentrations for which a quantitative value (10^1^ copies/reaction) was obtained. White circles surrounded with a dashed line indicate the relationship between the total number of inpatients and SARS-CoV-2 concentration including samples with concentrations below the limit of quantification (LOQ). Solid and dashed line indicate the regression line of the quantitative values and the including with below LOQ, respectively.

The concentrations in the samples taken on May 20, May 27, and June 3 of 2021, were LOQ or were not detected on three methods. The number of inpatients were large in these three days. The phenomenon indicates an inconsistency in the relationship between the concentrations and the number of inpatients. In order to interpret this inconsistency, the viral shedding period of the patients and the severity of the symptom need to be considered. It has been reported that the viral shedding period in feces from SARS-CoV-2 patients is up to 5 weeks, with an average of 20 days to 1 month (Wolfel et al., 2020, Xing et al., 2020, Wu et al., 2020, Xiao et al., 2020). According to Hu et al. (2020), most patients begin shedding from 3 days before to 3 days after the onset date, and it is estimated that the viral shedding begins on the day of onset in most cases. The peak value of viral shedding in feces often appears around 9 days after the onset (Wolfel et al., 2020). That means that the amount of virus released into feces from long-term inpatients is considered to be small, and it is necessary to extract patients who contribute to the amount of virus released into feces (i.e., the amount of virus in wastewater). In addition, in the severity determination of coronavirus infection patients in Japan by symptom, the severe and moderate patients thought to have difficult to walk independently and could not use the ward toilet. Thus, severe and moderate ill patients were excluded from the subjects, and only mild and asymptomatic patients were extracted as patients possibly contributing to the concentration in wastewater. The relationship between the number of inpatients and the SARS-CoV-2 concentration in wastewater is analyzed considering the number of days after onset and symptoms. Among mild ill and asymptomatic patients, the period of the virus shedding in the feces after the onset was assumed as some days, and only inpatients included in the number of days were extracted. In this study, the average number of days from onset to hospitalization was 4.5 days, and the average length of hospitalization until discharge was 17 days. Therefore, the extraction period was assumed to be from 5 to 20 days after onset and analyzed in order with 1-day increments. Furthermore, the below LOQ data in the virus concentration gives information that the concentration was low, although it was not quantitative. These obtained data were used as the assumed value for LOQ data in analyzing the relationship between the virus concentration and the number of patients considered with severity and days of after onset.

Fig. 4 shows the correlation between the virus concentration and the number of patients within the assumed period of the virus shedding after the onset. The correlation was highest in DC-MW, PEG-MW and PEG QVR when 7 days is assumed as the period of the virus shedding after the onset. Considering the existing report that the peak value of viral shedding in feces often appears around 9 days after the onset (Wolfel et al., 2020), it is reasonable that the highest correlation was observed when extracting the patients within 7 days after the onset. The results of this analysis suggest that when evaluating the correlation between viral concentrations in wastewater and the number of patients, it is necessary to consider the patient’s situation, i.e., the severity of symptoms and the number of days since onset. It was also expected that similar considerations should be applied to the risk management, such as estimating the number of patients, which is an important data use in WBE. The correlation between the patient’s contribution value to SARS-CoV2 load in feces and the SARS-CoV-2 concentration in wastewater from infectious diseases ward is shown in Fig. 5. The patient’s contribution value to virus load, which means the given additional weight to value of shedding more virus in the feces, was determined. A quadratic approximation curve was calculated for the fecal viral concentration data from patient for each day post-onset (Wolfel et al., 2020), and a contribution value was fitted to each patient. Compared to the relationship between the concentrations and the total inpatients (Fig.3), higher correlation was obtained. Therefore, if the severity of the patient’s symptoms and the number of days with virus shedding after the onset of the disease were taken into consideration, it is interpretable that the obtained values were smaller than the LOQ despite the large number of total inpatients.

**Fig.4.**
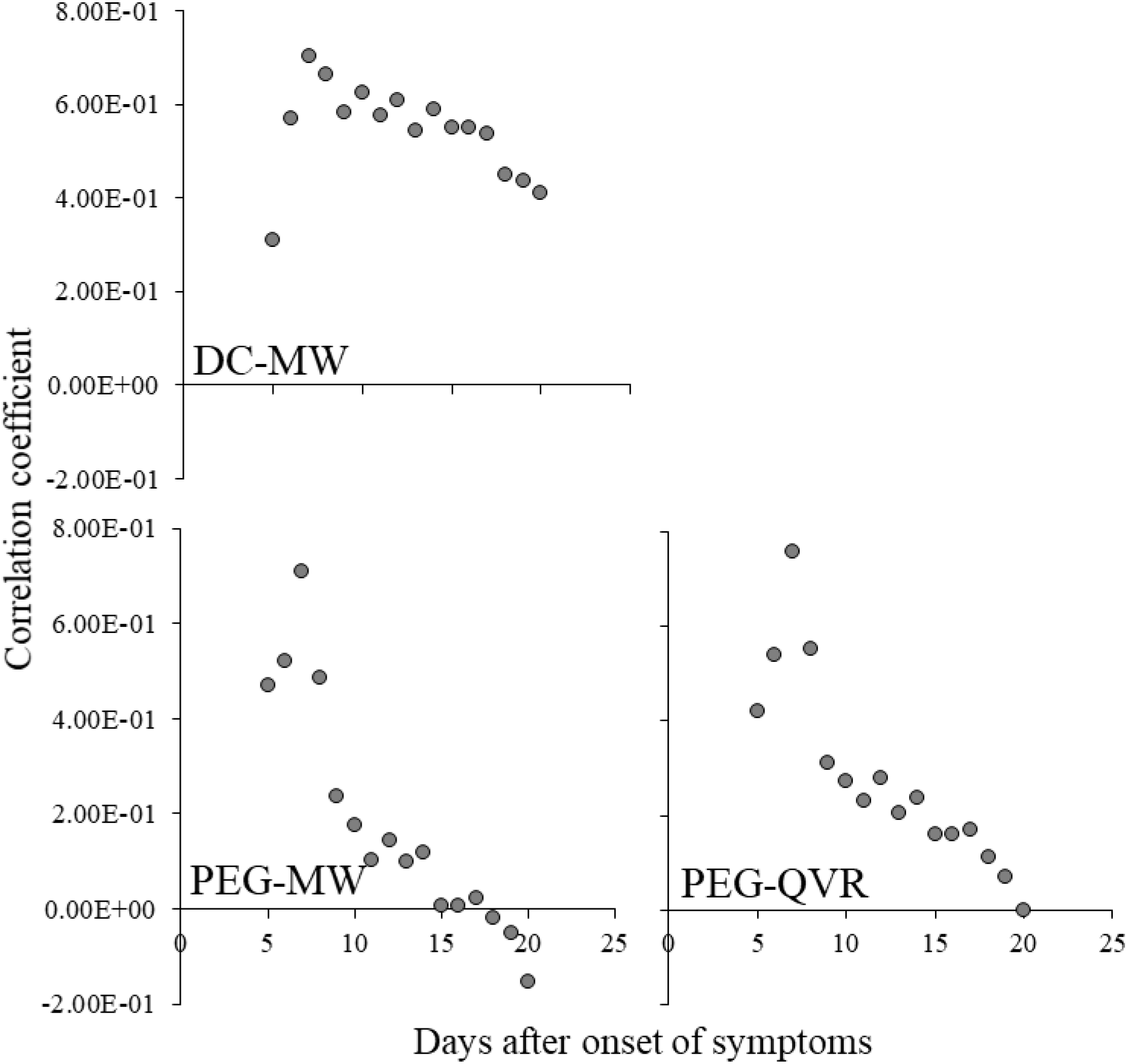
Correlation coefficient between SARS-CoV-2 concentration in wastewater form infectious diseases ward and patient number of days after onset of symptoms.

**Fig.5.**
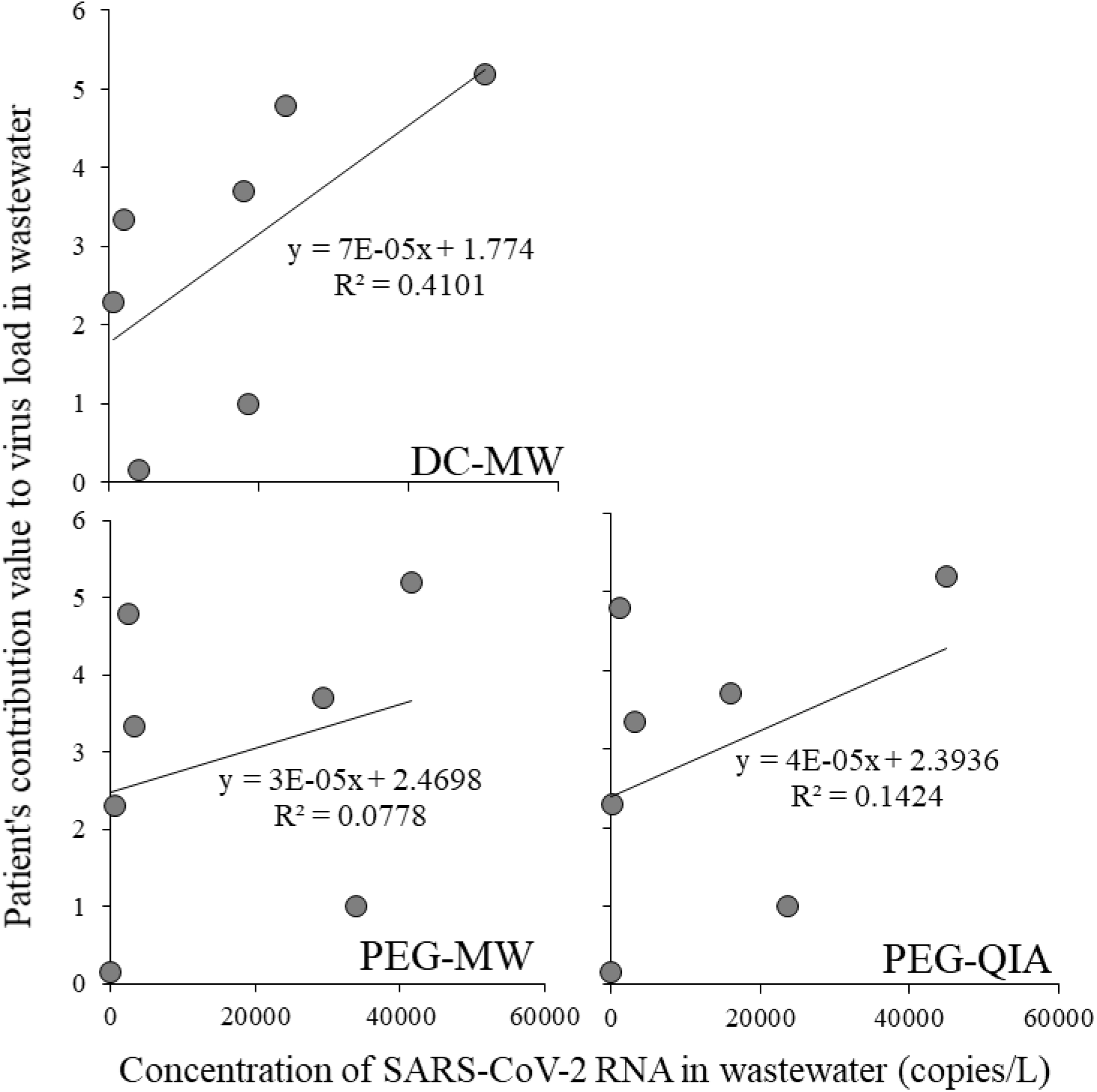
The correlation between the patient’s contribution values to SARS-CoV-2 load in feces and SARS-CoV-2 concentrations in wastewater from the infectious diseases ward.

According to the previous studies, the virus can be released into feces for a long period of time (Wolfel et al., 2020; Wu et al., 2000, Xiao et al., 2020, Xing et al., 2020). Because the amount of virus shedding is expected to decrease, the correlation with the virus concentration in the wastewater will be low when long-term inpatients are included; however, it is considered that a certain extent of correlation remains. Thus, the strength of the correlation will be gradually weakened as the assumed period of the virus shedding after the onset increases. Among the three conditions, only DC-MW showed the above-mentioned trend while the other two conditions showed a rapid drop of the correlation coefficient (around 9 days after onset). These suggest that the result of DC-MW is more realistic, and DC-MW has the high sensitivity and quantitatively in the actual field. Therefore, DC-MW is considered to be superior to the other two methods in monitoring the actual situation.

## Conclusion

In this study, we compared the methods of concentration and extraction-purification of the viruses in wastewater. By comparing the efficiency of the methods with the addition of process control, DC-MW was confirmed to be the most sensitive and efficient method. The result of quantification and detection of the actual SARS-CoV2 in the wastewater, the positive result was confirmed even in the sample which could not be detected by other methods, and was shown also highly sensitive. In addition, regarding the correlation between the number of patients and the virus concentration in the wastewater, the DC-MW was found to be highly sensitive and efficient, and was likely to be advantageous in reflecting the actual situation. Because this method targets the genome, the infectious viruses cannot be evaluated. For enteric viruses such as rotavirus, to recover the infectious viruses is necessary and important for assessment of the infection risk in environment and the accumulation in marine products at the discharged area. In such cases, this method may not be suitable. On the other hand, for the case of SARS-CoV-2, the main target on WBE is to detect earlier the patients exist and the epidemic in the collection area or the building of interest, indicating that the genome detection meets the requirements.

From another point of view, the risk of infection from wastewater is very low for the case of SARS-CoV-2, but it is not negligible for the case of norovirus. Because the virus inactivation process is included in the first stage of DC, the safety of workers is high. In particular, when the effluent standards for viruses in wastewater are established in the future, workers who are not accustomed to handling viruses will be able to conduct the sampling and quantification. The high safety is an advantage in such cases. This method is highly safe and simple method that can reduce the influence of the operator’s technique. The highly sensitive, rapid and simple method of DC-MW is also considered to be effective for the stable data accumulation.

## Data Availability

All data produced in the present study are available upon reasonable request to the authors

## ACKNOWLEDGEMENT

This work was supported by JSPS KAKENHI Grant Number 20H00632.

